# Understanding Patients Experiences Living with Diabetes Mellitus: A Qualitative Study, Gujarat, India

**DOI:** 10.1101/2021.12.06.21267307

**Authors:** Angel Christian, Kailash Nagar

## Abstract

**Introduction:** Diabetes mellitus is leading health problem in India. India is experiencing the burden of communicable disease as well as non-communicable disease. It is believed due to unhealthy life style and faulty food habits. The International Diabetes Federation estimated that 72.9 million adults in India were living with diabetes in last four years. The study overviews the different factors like personal, familial, social, and financial affecting the life style and management of the disease. Thus, diabetes mellitus affects all the dimensions of health of an individual. The aims of the study to assess quality of life and experiences faced by the patients who are suffering from diabetes mellitus and how the progressive stage of the diseases altered the all the dimensions.

**Objectives:**

1. To explore the lived in experiences of diabetes mellitus patients.
2. To explore the various dimensions of the lived in experiences of diabetes mellitus patients.

**Materials and Methods:** A qualitative, exploratory and descriptive design was used to explore and describe the experiences lived by participants suffering from diabetes in the selected areas of Kheda district Gujarat. The investigator used the phenomenological approach of the qualitative paradigm. The study conducted among 10 samples who are suffering from Diabetes Mellitus more than 07 years, sampling technique used was non-probability. Semi structured interview questions were used to conduct the detail history from the participants, where investigator has attain at least 6-8 sitting, (duration 30-45 Min.) with each participants.The investigator used local language for the interview and recorded in mobile, later all the recorded session has been verbatimaccording to the themes and sub-themes.

**Results:** The study resulted in the extraction of six themes, which reflect the experiences of the study participants. The themes are: 1) lived time; 2) lived body; 3) lived relationship; 4) lived economical; 5) lived vocational and 6) lived spiritual. It was found that diabetes still causes participants to suffer from fear, depression and anxiety along with they hate to have modification in dietary pattern in which they cannot have sweets and sweet products. Although emotional support was provided by the family members and friends. Participant’s absence from work in their early diagnosis was due to both their health conditions and emotional embarrassment. There are evidences that diabetes has psychological damage to patient’s life and so they feel more anxious and fatigue.

**Conclusion:** The study attempted to explore the lived in experiences of Diabetes Mellitus Patients and it was found that DM has affected all the dimensions of health especially in physical, psychological and economical dimensions. 1) In Physical dimension the patients were having common complaints of giddiness, weakness and headache which was leading them to certain problems. 2) In psychological dimension the patients were having more anxiety, fear regarding the disease and long term treatment along with depression. 3) In economical dimension some patients were having good family income so there were no issues related to expenses on diet and medication 4) In social dimension all the participants were having good relation with their everyone. 5) In vocational dimension all were having difficulty at work place to concentrate in their work but after starting the treatment their health was improved so later they continued their work properly. 6) In spiritual dimension only two participants were not having belief in God rest all were having faith in God and prayed daily.

## Introduction

Diabetes mellitus is a persistent illness that happens when the pancreas doesn’t create[1] appropriate insulin or the body don’t utilize the insulin delivered. Insulin is a chemical that directs glucose[2].

Excessive blood sugar or hyperglycemia[3], is a common effect of not controlling diabetes[4]and in that time leads to serious damage[5] to many of the body’s systems, especially the nervous system and the blood vessels[6].

As of now, India is viewed as the ‘Diabetes Capital’ of the world[7]. This is on the grounds that the biggest number of individuals with diabetes lives in this country[8]. Asian Indians have a higher inclination to insulin obstruction, diabetes mellitus and coronary vein infection[8-9]. In this milieu of high predominance of diabetes in India[10], critical exploration ends up working on the nature of care for the patients[9].

As indicated by the Diabetes Atlas 2006 distributed by the International Diabetes Federation[11], the quantity of individuals with diabetes in India at present around 40.9 million is relied upon to ascend to 69.9 million by 2025[12] except if earnest preventive advances are taken[12-13]. The supposed “Asian Indian Phenotype” alludes to specific remarkable clinical and biochemical irregularities in Indians which incorporate expanded insulin obstruction, more noteworthy stomach adiposity[14].

Diabetes mellitus is in reality described on the premise of hyperglycemia. It is, however, an extraordinarily heterogeneous sickness[14]. A primary increase turned into made inside the late Sixties while insulin-structured diabetes mellitus (type1) turned into prominent from non-insulin-established diabetes mellitus (type 2)[15].

## MATERIALS AND METHODS

### Research Design

A Qualitative research design

### Sampling methods

Non probability sampling method was used in the study

## INSTRUMENT FOR DATA COLLECTION

### Study samples

The sample size were 10 participants

### Study setting

Rural Area of Kheda District Gujarat

### Inclusion Criteria

1. Patients who have diabetes mellitus more than 07 years.
2. Patients who have diabetic foot ulcer (with more than 07 year DM).
3. Both male and female patients.
4. DM patients whose age is between 30-70 years.

## DATA COLLECTION

### Phase I

The investigator made the diabetes patient comfortable and maintained good rapport with them. Then the investigator asked questions regarding their lived-in experiences until the investigator achieved the saturation of data. The statements expressed by the patients were recorded in a mobile recorder. Saturation was accomplished through a triangulation of different data collection techniques, periods, places and participants. The time spent for each interview was approximately 30-45 minutes and minimum 6 to 10 sittings with each participant.

### Phase II

The audio taped interview was listened and transcribed into verbatim on same day. The collected data was analyzed using Colazzi’s analysis.

## DATA ANALYSIS

The data was analyzed using descriptive statistics.

### Modified Colizzi’s Analysis Framework

1. All interview was deciphered into word for word and read to get a comprehension into word for word.
2. Significant picked phrases identifying with the experience of patient being examined were eliminated.
3. Significant statements were organized into cluster of themes.
4. The themes were used to provide the full description of the experiences.

## RESULTS

**Table 1** reveals the distribution of demographic variables among 10 samples (patients with diabetes mellitus). In terms of age 40% (4) participants belongs to the age group of 51-60 and last 20% (2) belongs to 61-70 years.In regard to gender differentiation 80% (8) of the participants were male. Out of 100%, 30% (3) of the participants were Christian and rest 70% (7) participants were Hindu. Regarding the educational qualification, 40% (4) participants had done secondary education, In regard to the occupational status, 50% (5) of the participants were private Employees.

**Table 1:** Frequency and percentage distribution of diabetes patients according to age, gender, educational qualification, occupation, family income per month and religion. n=10

| VARIABLE | CLASSIFICATION | FREQUENCY | % |
| --- | --- | --- | --- |
| Age in years | a) 30-40 | 2 | 20% |
|  | b) 41-50 | 2 | 20% |
|  | c) 51-60 | 4 | 40% |
|  | d) 61-70 | 2 | 20% |
| Gender | a) Male | 8 | 80% |
|  | b) Female | 2 | 20% |
|  | c) Transgender | - | - |
| Religion | a) Hindu | 7 | 70% |
|  | b) Christian | 3 | 30% |
|  | c) Muslim | - | - |
|  | d) Others | - | - |
| Educational background | a) No formal Education | - | - |
|  | b) Primary Education | - | - |
|  | c) Secondary Education | 4 | 40% |
|  | d) Higher Secondary | 3 | 30% |
|  | e) Degree | 3 | 30% |
| Occupation | a) House wife | - | - |
|  | b) Government job | 2 | 20% |
|  | c) Private | 5 | 50% |
|  | e) Business | - | - |
|  | f) Others | 3 | 30% |
| Diet | a) Vegetarian | 2 | 20% |
|  | b) Non- vegetarian | - | - |
|  | c) Mixed | 8 | 80% |

**Table 2** reveals the distribution of demographic variables among 10 samples (patients with diabetes). Out of which 20% (2) were unmarried and rest 80% (8) were married.

**Table 2:** Frequency and percentage distribution of Diabetes Mellitus Patients according to the type of family, history of diabetes, type of diabetes and treatment category. n=10

| VARIABLE | CLASSIFICATION | FREQUENCY | % |
| --- | --- | --- | --- |
| Marital status | a) Single | 2 | 20% |
|  | b) Married | 8 | 80% |
|  | c) Divorce | - | - |
|  | d) Widower/widow | - | - |
| Family type | a) Nuclear | 6 | 60% |
|  | b) Joint | 4 | 40% |
|  | c) Extended | - | - |
| Family history of DM | a) yes | 4 | 40% |
|  | b) No | 6 | 60% |
| Type of DM | a) Type I | 1 | 10% |
|  | b) Type II | 9 | 90% |
| Any medication taken | a) Yes | 10 | 100% |
|  | b) No | - | - |
| Duration of DM | a) 8 to 10 years | 7 | 70% |
|  | b) 11 to 13 years | 3 | 30% |
|  | c) 14 to 16 years | - | - |
|  | d) 17 years above | - | - |

Out of the 10 participants 60% (6) belonged to nuclear family and 40% (4) belonged to joint family.In regards to family history of diabetes, 60% (6) of the participants were not having family history of diabetes and 40% (4) of them had family history of diabetes.90% (9) participants belongs to type II diabetes mellitus. Duration of suffering from diabetes in which70% (7) participants were suffering from 8-10 years and 30% (3) participants were suffering from 11-13 years from diabetes.

## DIMENSIONS USED FOR DATA COLLECTION

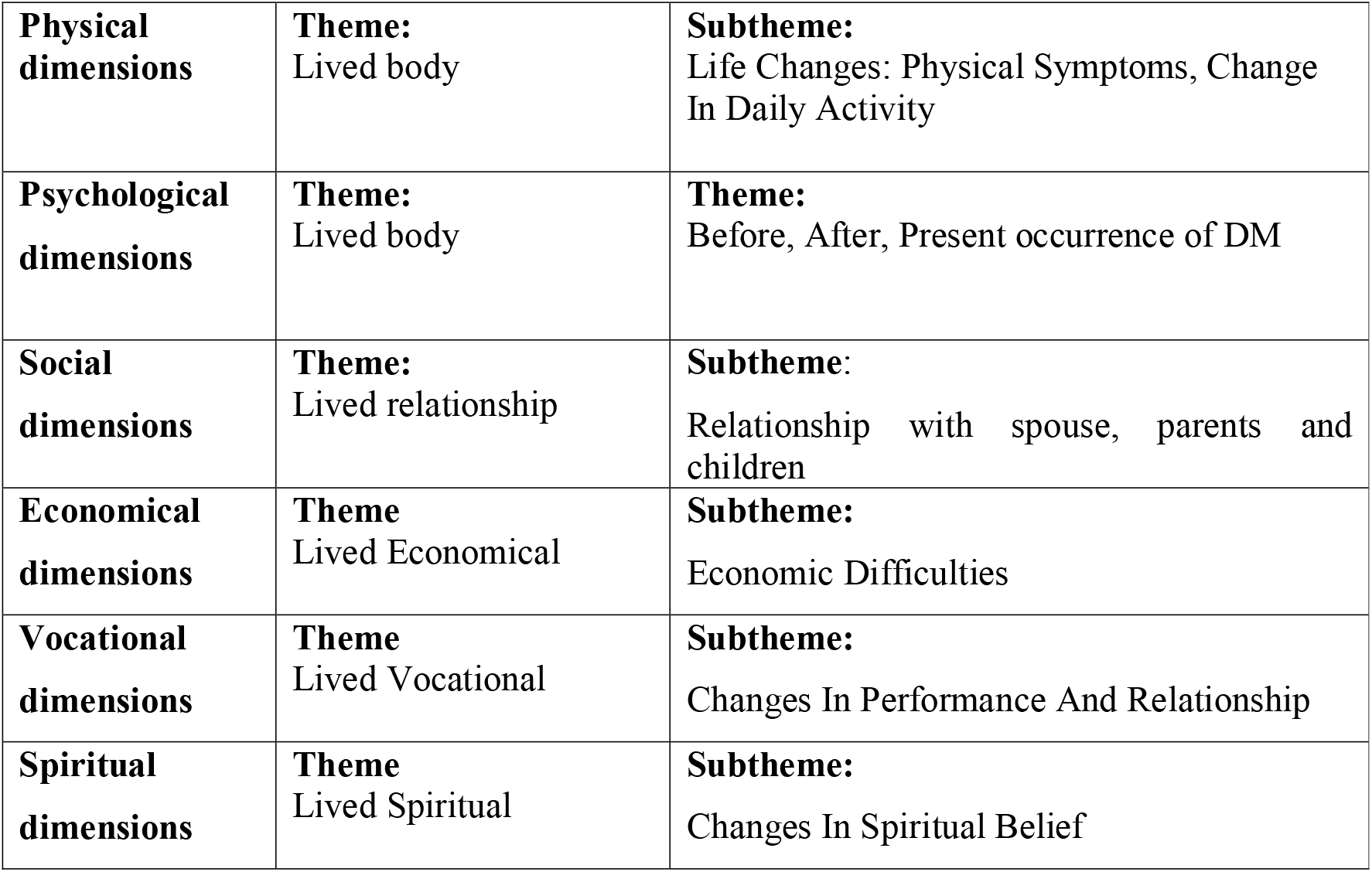

## DISCUSSION

The objective of the study was to explore the lived-in experiences of diabetes patients and to explore the various dimensions of lived-in experiences of diabetes patients.

### Theme 1: Lived Time (before and after being diagnosed as having DM.)

#### Subtheme 1: Before: Denial and difficulty to accept the disease condition

All my participants are having same review that *“why this disease have occurred to me, this should not have happen.”*

#### Subtheme 2: After: Acceptance of the disease condition

Most of all my participants are having review *that “now what to do, it has already happened now I will take medicine.”*

#### Subtheme 3: Present: Uncertainty regarding future

All my participants are having fear regarding the long term treatment and don’t like to take medicines for long time and wants to get well soon.

### Theme 2: Lived Body

#### Subtheme 1: Life changes: Physical Symptoms

Some of my patients were having headache, giddiness and nausea, two clients were having body pain and only one had vomiting.

#### Subtheme 2: Life changes: Change in daily activities

All my clients are were loving to eat sweets and spicy foods but due to diabetes mellitus they were having restrictions and unable to eat all good sweets and even diet plan was changed so they were very nervous about their changes in all.

### Theme 3: Lived Relationship

#### Subtheme 1: Relationship with wife/husband

Seven of my patients were married and their wife/husband were all supportive and taking good care of them.

#### Subtheme 2: Relationship with parents

All my clients were having good relationship with their parents and they all supported them very well.

#### Subtheme 3: Relationship with children

Seven clients were married and their all children were very helpful and supportive to them and even advised them about the medicines and diet.

#### Subtheme 4: Relationship with siblings

Few siblings were supportive to some participants and to other participants the siblings were not bothered.

### Theme 4: Lived economical

#### Subtheme 1: Economical difficulties

Only three participants were having financial problems rest all participants were stable economically.

#### Subtheme 2: Increased expenditure: Difficulties in meeting family needs

Three participants’ families were having crises sometimes due to more expenses. They were taking their medicines from the PHC and so medical problems were not there but problems regarding to diet and daily living.

#### Subtheme 3: Family members had to work: to manage financial difficulties

None of the families were having that much financial problems that after being diagnosed as DM their other family members had to work.

#### Subtheme 4: Reactions on the expenses on the treatment: Positive

Five participants were taking medicines from PHC and others were taking medicines from private dispensary as they were financially healthy.

### Theme 5: Lived Vocational

#### Subtheme 1: Changes in performance: Low standard of performance

All my participants were having problems in their work as they were having physical symptoms and not well due to body pain and weakness along with fatigue so till they all were not diagnosed and not started the treatment their performance was not good but later on it came to normal.

#### Subtheme 2: Change in relationship with colleagues

All the colleagues of participants were supportive and cooperative at the time of need during the illness faced.

### Theme 6: Lived spiritual

#### Subtheme 1: Change in spiritual belief: Change in belief toward God

All my participants were having faith in God except one and they prayed to God daily for the healing that they get well soon.

#### Subtheme 2: Spiritual relation: High intensified belief towards God

All participants were having belief that God can only heal them and only one participant was unfaithful and do not believe in God.

## MAJOR FINDING ACCORDING TO THEMES

The finding of the study showed that being diagnosed as diabetes brings fear, tensions and frustration among the patients. The participants also faced the problems in controlling the diet as most of all participants liked to eat sweets and rice. They were having difficulty in the functions to attend as all the foods were having delicious sweets and spicy foods. Participants also told that family support is the most important part in which they feel cared and supported by them. Diabetes also affected the participants psychologically as they had to take long term treatment and need to continue regularly.The study also revealed that it also affects the patients vocationally as they felt more tiredness and weakness due to their work load and sometimes stress may also affect. Some participants those who were taking treatment on private basis also affect their financial condition so they sometimes took help from their siblings and relatives.

## Data Availability

yes all data has been mentioned in the manuscript such as abstract aims objective methodology result and conclusion

## ETHICAL CONSIDERATION

The study was approved by the institutional ethical committee of Dinsha Patel College of nursing, research committee, there are total 15 members in the committee from various field. This study was approved by Institutional Ethics Committee of Dpcn.2020-21, Proposal ID: DPCN-IEC//2020-21/08 and a formal written permission was gathered from the authority of or Principal of Instituteprior to data collection.

